# Cerebrospinal Fluid from Cytology Supernatant as a Liquid Biopsy Specimen for Diagnosis and Surveillance for Pediatric Primary Central Nervous System Tumors

**DOI:** 10.1101/2025.09.30.25336820

**Authors:** Nicholas Chun, Brandon Edelbach, Muhammad Baig, Laura A. T. Kagami, Richard A. Robison, Edwina Choung, Isaac Kremsky, Kevin Leeper, Brian Ferguson, D. Gigi Ostrow, Cindy Fong, Udochukwu Oyoyo, Rajeev Nowrangi, Minwoo Song, Bin Othman, Tanya Minasian, Diem Kieu Tran, Sujata Mushrif, Adina Achiriloaie, Saied Mirshahidi, Gary Yu, Pramil Singh, Ravi Raghavan, Jaclyn A. Biegel, Yan Chen Wongworawat

## Abstract

**Background:** Assessing circulating cell-free DNA (cfDNA) in cerebrospinal fluid (CSF) has been proposed as a promising alternative to tissue biopsy. Advances in cfDNA sequencing have further underscored the potential of CSF liquid biopsies. CSF is routinely collected for cytologic evaluation at diagnosis, and at recurrence in both pediatric and adult central nervous system (CNS) tumors.

**Methods:** As part of a pilot study, CSF specimens were prospectively collected from seven pediatric patients with primary CNS malignant tumors. When possible, CSF was collected fresh and/or from processed cytology specimens. Low-pass whole genome sequencing (LP-WGS) and next-generation sequencing (NGS) using a custom targeted sequencing panel were performed on the specimens to identify copy number alterations (CNAs), detect mutations, and estimate circulating tumor DNA (ctDNA) fractions. Results were compared with matched tumor tissue molecular profiles and corresponding imaging findings.

**Results:** Abnormalities in cfDNA were detected in four patients. Sequencing of CSF cytology supernatants demonstrated the presence of circulating tumor DNA with characteristic CNAs and mutations that matched what was seen the tumor tissue as well as the fresh CSF specimens. These studies also revealed tumor heterogeneity and genomic evolution over time.

**Conclusion:** This study demonstrates the feasibility of utilizing routinely discarded supernatants from CSF cytology specimens for LP-WGS and targeted NGS. Our approach optimizes the use of CSF that may be limited in pediatric patients as a reliable source for liquid biopsy-based genomic studies. Future research will be necessary to optimize the methodology to enable clinical implementation.

**Key points:**

- Combined low-pass whole genome sequencing (LP-WGS) and targeted next-generation sequencing (NGS) can detect circulating tumor DNA (ctDNA) in cerebrospinal fluid (CSF) cytology supernatant samples.
- Our approach transforms clinically discarded specimens into a valuable and reliable source for liquid biopsy.
- LP-WGS and targeted NGS of CSF cytology-derived cell-free DNA (cfDNA) is more sensitive than CSF cytology for tumor detection at initial diagnosis and during surveillance.

**Importance of Study:** This report highlights the feasibility of utilizing supernatants from cerebrospinal fluid (CSF) cytology specimens that would normally be discarded for diagnosis and follow up for patients with pediatric primary central nervous system (CNS) tumors, using the LBSeq4Kids combined low-pass whole genome sequencing (LP-WGS) and targeted next-generation sequencing (NGS) liquid biopsy platform. It optimizes the use of limited clinical specimens and maximizes diagnostic and research potential, yet does not pose additional risk to patients by utilizing routinely collected cytology specimens. Identification of copy number alterations (CNAs) and DNA sequence variants detected in CSF-derived cell-free DNA from patients with high grade glioma and embryonal tumors can augment tissue-derived molecular analyses. CSF liquid biopsy approaches have the potential to transform clinical practice for improved diagnosis, risk stratification, and disease surveillance.

## Introduction

Liquid biopsy utilizing cerebrospinal fluid (CSF)-derived circulating cell-free DNA (cfDNA) has been proposed as a promising alternative to tissue biopsy. Unlike plasma-derived circulating tumor DNA (ctDNA), which is often minimal or absent due to the limitations imposed by the blood-brain barrier, CSF is in direct contact with most malignant central nervous system (CNS) tumors, resulting in an enrichment of tumor-derived ctDNA^1^. Moreover, the paucicellular nature of CSF reduces the background noise from non-tumor DNA, enhancing the sensitivity of molecular analyses^2^. Tumors located in the brainstem, ventricular, periventricular, and sub-pial/sub-arachnoid regions, which are in close proximity to CSF, are therefore ideal candidates for this approach^3^.

Advances in cfDNA sequencing have further underscored the potential of CSF liquid biopsy. Highly sensitive detection methods—ranging from targeted approaches such as digital droplet PCR (ddPCR) and beads, emulsion, amplification, magnetics (BEAMing) PCR to broad next-generation sequencing (NGS) panels have successfully identified tumor-specific mutations and copy number alterations (CNAs) in CSF-derived cfDNA. For instance, Pan et al. identified tumor-specific mutations in over 82.5% of brainstem tumor cases using a 68-gene NGS panel, while Panditharatna et al. demonstrated that ddPCR detection of the *H3K27M* mutation can serve as an effective surveillance tool in pediatric diffuse midline gliomas^4,5^. O’Halloran *et al*. proposed a more comprehensive approach employing LP-WGS and a targeted NGS gene panel, which detected genome-wide CNAs, mutations, and gene fusions^6^. In addition, longitudinal CSF liquid biopsy studies, as reported by Miller *et al*., have revealed dynamic changes in growth factor receptor signaling pathways throughout the disease course, highlighting its utility in tracking tumor evolution^7^.

CSF samples are routinely collected in this setting for cytologic evaluation at primary diagnosis, and at recurrence in selected pediatric primary malignant CNS tumors. While there is a rapidly developing body of literature surrounding the utilization of CSF-derived cfDNA for the diagnosis and possible monitoring of treatment response, these studies rely on fresh CSF^1-11^. Although the volume of CSF obtained in such scenarios is often sufficient for both cfDNA and cytologic analysis in adults, this issue is particularly relevant in pediatric patients, who generally have smaller body size and lower CSF volumes. Standard preparation of CSF for cytology involves centrifugation, after which the cell pellet is transferred to slides for microscopic evaluation, while the supernatant is typically discarded. To date, no one has reported the possible benefits of using ctDNA obtained from the discarded CSF cytology supernatant for diagnosis and ongoing patient monitoring. In this study, we prospectively enrolled pediatric patients at diagnosis and at recurrence to evaluate the utility of CSF cytology supernatant-derived cfDNA for the diagnosis, characterization and follow-up of primary malignant CNS tumors, and to investigate whether the cytology supernatant can serve as a valid source for cfDNA sequencing.

## Materials and Methods

### Patient recruitment and data acquisition

The study was approved by the Loma Linda University (LLU) Institutional Review Board (IRB # 5240377). All patients were recruited from LLU. Informed consent was obtained for each patient by qualified study personnel in accordance with the Declaration of Helsinki. Children were provided with age-appropriate information about the study and given the opportunity to express their willingness to participate. Inclusion criteria were as follows: (1) individuals less than 18 years of age, (2) who had been diagnosed with a primary CNS tumor and treated at LLU, and (3) were in stable condition to undergo a lumbar puncture or other method of CSF collection per standard of care. Patient demographics, diagnosis, radiographic imaging, and treatment schedules were gathered from patient records. Basic demographic information, pathology diagnosis, together with molecular alterations from tumor tissue and CSF liquid biopsy, are shown in **Table 1**. Specimens from cases 7, 9, and 10 were collected for future studies.

**Table 1.**
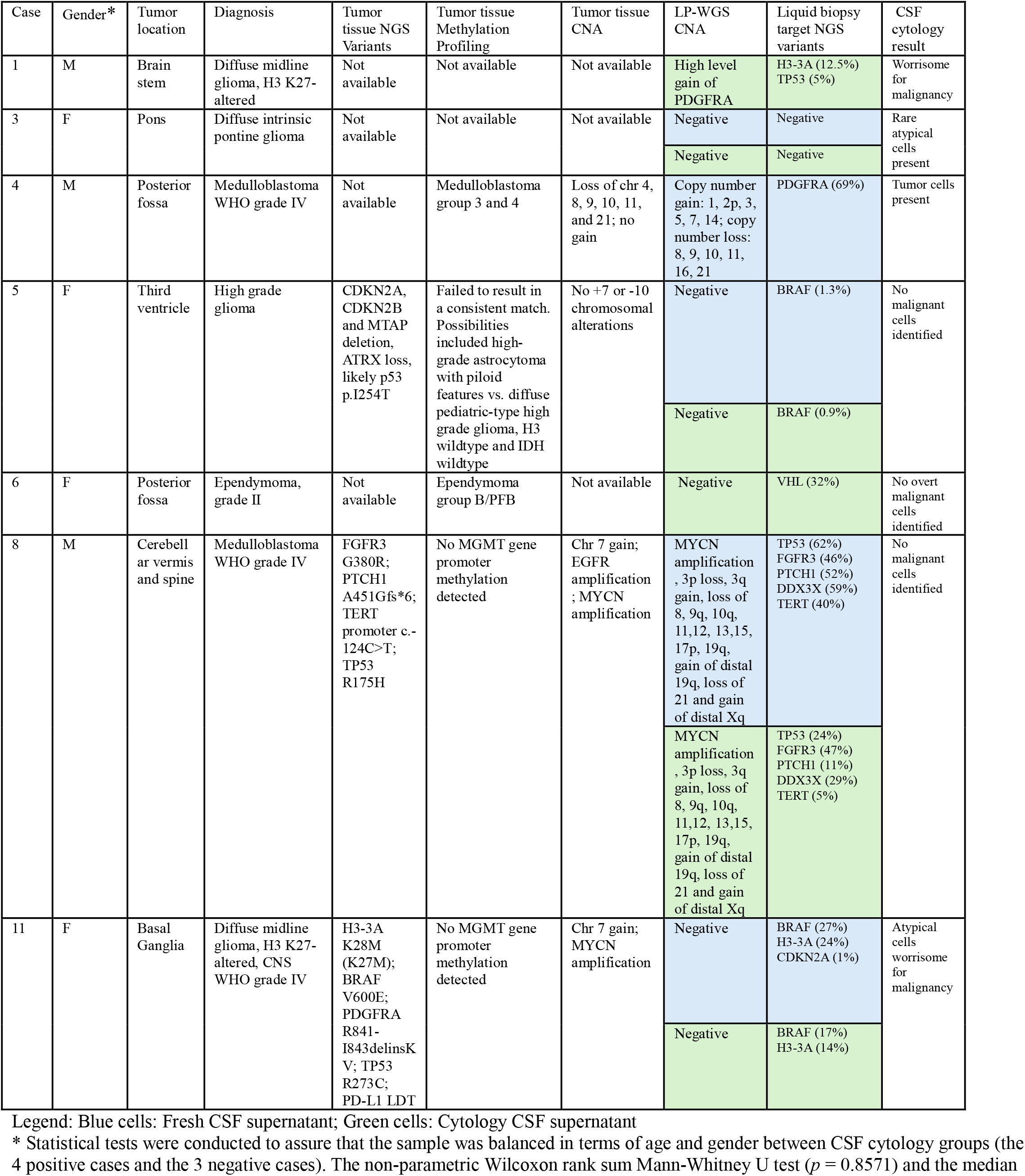
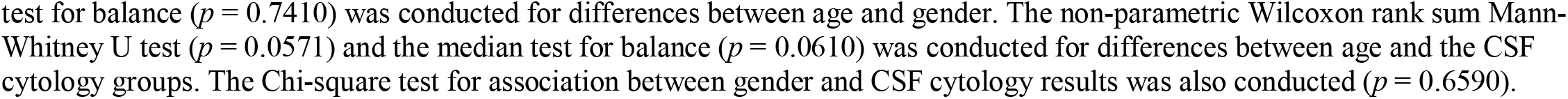
CNAs and genetic mutations identified by LP-WGS and targeted NGS.

| Case | Gender* | Tumor location | Diagnosis | Tumor tissue NGS Variants | Tumor tissue Methylation Profiling | Tumor tissue CNA | LP-WGS CNA | Liquid biopsy target NGS variants | CSF cytology result |
| --- | --- | --- | --- | --- | --- | --- | --- | --- | --- |
| 1 | M | Brain stem | Diffuse midline glioma, H3 K27-altered | Not available | Not available | Not available | High level gain of PDGFRA | H3-3A (12.5%) TP53 (5%) | Worrisome for malignancy |
| 3 | F | Pons | Diffuse intrinsic pontine glioma | Not available | Not available | Not available | Negative | Negative | Rare atypical cells present |
|  |  |  |  |  |  |  | Negative | Negative |  |
| 4 | M | Posterior fossa | Medulloblastoma WHO grade IV | Not available | Medulloblastoma group 3 and 4 | Loss of chr 4, 8, 9, 10, 11, and 21; no gain | Copy number gain: 1, 2p, 3, 5, 7, 14; copy number loss: 8, 9, 10, 11, 16, 21 | PDGFRA (69%) | Tumor cells present |
| 5 | F | Third ventricle | High grade glioma | CDKN2A, CDKN2B and MTAP deletion, ATRX loss, likely p53 p.I254T | Failed to result in a consistent match. Possibilities included high-grade astrocytoma with piloid features vs. diffuse pediatric-type high grade glioma, H3 wildtype and IDH wildtype | No +7 or -10 chromosomal alterations | Negative | BRAF (1.3%) | No malignant cells identified |
|  |  |  |  |  |  |  | Negative | BRAF (0.9%) |  |
| 6 | F | Posterior fossa | Ependymoma, grade II | Not available | Ependymoma group B/PFB | Not available | Negative | VHL (32%) | No overt malignant cells identified |
| 8 | M | Cerebellar vermis and spine | Medulloblastoma WHO grade IV | FGFR3 G380R; PTCH1 A451Gfs*6; TERT promoter c.-124C>T; TP53 R175H | No MGMT gene promoter methylation detected | Chr 7 gain; EGFR amplification; MYCN amplification | MYCN amplification, 3p loss, 3q gain, loss of 8, 9q, 10q, 11,12, 13,15, 17p, 19q, gain of distal 19q, loss of 21 and gain of distal Xq | TP53 (62%) FGFR3 (46%) PTCH1 (52%) DDX3X (59%) TERT (40%) | No malignant cells identified |
|  |  |  |  |  |  |  | MYCN amplification, 3p loss, 3q gain, loss of 8, 9q, 10q, 11,12, 13,15, 17p, 19q, gain of distal 19q, loss of 21 and gain of distal Xq | TP53 (24%) FGFR3 (47%) PTCH1 (11%) DDX3X (29%) TERT (5%) |  |
| 11 | F | Basal Ganglia | Diffuse midline glioma, H3 K27-altered, CNS WHO grade IV | H3-3A K28M (K27M); BRAF V600E; PDGFRA R841-I843delinsKV; TP53 R273C; PD-L1 LDT | No MGMT gene promoter methylation detected | Chr 7 gain; MYCN amplification | Negative | BRAF (27%) H3-3A (24%) CDKN2A (1%) | Atypical cells worrisome for malignancy |
|  |  |  |  |  |  |  | Negative | BRAF (17%) H3-3A (14%) |  |

Legend: Blue cells: Fresh CSF supernatant; Green cells: Cytology CSF supernatant
\* Statistical tests were conducted to assure that the sample was balanced in terms of age and gender between CSF cytology groups (the 4 positive cases and the 3 negative cases). The non-parametric Wilcoxon rank sum Mann-Whitney U test ( $p = 0.8571$ ) and the median
test for balance ( $p = 0.7410$ ) was conducted for differences between age and gender. The non-parametric Wilcoxon rank sum Mann-Whitney U test ( $p = 0.0571$ ) and the median test for balance ( $p = 0.0610$ ) was conducted for differences between age and the CSF cytology groups. The Chi-square test for association between gender and CSF cytology results was also conducted ( $p = 0.6590$ ).

### Sample Collection

CSF samples were obtained through standard of care procedures such as lumbar puncture (LP), ventricular-peritoneal (VP) shunt taps, external ventricular drain (EVD) and intra-operatively. Approximately 4-8ml of CSF was collected. If CSF volume was sufficient, the CSF was divided into two parts: 1) a fresh specimen and 2) a preserved sample (in CytoLyt, Hologic Inc, Marlborough, MA) for routine cytologic examination. If the volume was insufficient, only the supernatant from the CSF cytology was processed. In cases where the CSF cytology specimen was unavailable, the fresh CSF was processed in a manner that simulated a typical cytology procedure: CytoLyt was added to the fresh CSF specimen in a 4:1 ratio (CSF:CytoLyt). The mixture was centrifuged to separate the supernatant from the cell pellets. The cell pellets were frozen immediately. The supernatant was held at 4°C for three days before being frozen at -80°C. The supernatant was considered a proxy for CSF cytology supernatants, as it captured the degradation effects of storage.

### Purification, Processing and Analysis of Specimen

The fresh CSF specimens were centrifugated at 3,000 x g at 4°C for 10 min to remove intact cells, and the supernatant was collected and stored at -80°C. For the preserved CSF from the cytology specimen the cells were transferred to slides for staining and microscopic evaluation, and the leftover supernatant, which is typically discarded, was frozen at -80°C. The specimens were then shipped to the Center for Personalized Medicine at the Children’s Hospital Los Angeles (CHLA) for cfDNA extraction, low-pass whole genome sequencing (LP-WGS) and targeted sequencing as previously described ^6,10^. cfDNA quantity and quality were assessed using the Agilent TapeStation High Sensitivity DNA D1000 assay (Santa Clara, CA) in accordance with established protocols ^6^. The currently validated LBSeq4Kids assay includes LP-WGS for copy number variant analysis and a custom targeted sequencing panel (TSP) (Twist Bioscience) that is used to sequence the full coding exons of 136 tumor associated genes as well as fusions involving *BRAF, FOXO1* and *EWSR1*. For LP-WGS, reads are binned into 500 kb regions across the entire genome and the ichor CNA algorithm is used for segmented calls and ctDNA fraction estimation in cfDNA. Annotation of the targeted panel was performed with VarSeq (Golden Helix) and verified using Integrative Genomics Viewer (IGV)^12^.

## Results

### cfDNA can be successfully isolated from the CSF cytology supernatants yielding a cfDNA content comparable to that obtained from fresh CSF

The proportion of cfDNA fragments in the 120–220 bp size range is correlated with cfDNA concentration in CSF. The points of comparison, CSF collection method, total cfDNA concentration, estimated circulating tumor DNA (ctDNA) fraction from iChor results, as well as days between collection and freezing are shown in **Supplementary Table 1**. For example, sample Case 3 includes matched fresh CSF and the cytology supernatant. The total DNA concentration in the CSF cytology supernatant is reduced compared to fresh CSF (2.8 ng/ul vs. 12.5 ng/ul), and the cfDNA concentration was also lower in CSF cytology supernatant (0.47 vs 1.63 ng/µL); however, the relative cfDNA% was higher (17% vs 13%) (**Fig. 1 A and B**). **Figure 1C** illustrates the total cfDNA concentration for fresh CSF vs CSF cytology supernatant for each case.

**Figure 1.**
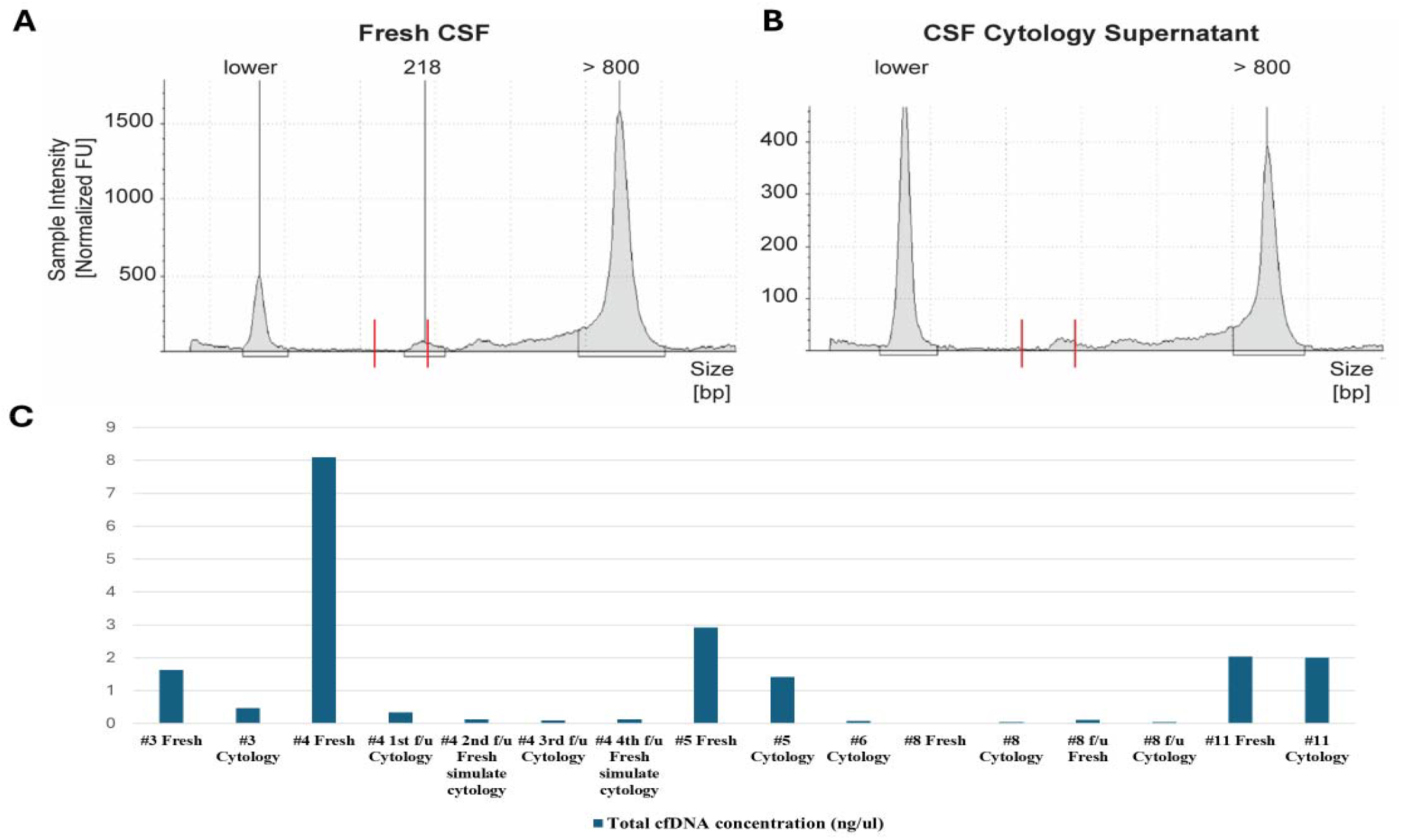
**(A-B)** The total DNA concentration from Case 3 in the CSF cytology supernatant **(B)** was lower than fresh CSF (**A**) (2.8 ng/ul vs. 12.5 ng/ul), and the total cell free DNA (cfDNA) concentration was also lower in CSF cytology supernatant (0.47 vs 1.63 ng/µL); however, the relative cfDNA% was higher (17% vs 13%). Red lines: cfDNA fragments size (120–220 bp). **(C)** The total cfDNA concentration for fresh CSF and/or CSF cytology supernatant in each case.

### CNAs and specific mutations can be successfully detected through cfDNA sequencing from the cytology supernatants

The CNAs and mutations identified by LP-WGS and targeted NGS from fresh CSF, CSF cytology supernatant and tumor tissue are shown in **Table 1**.

Representative findings are discussed in the case examples below.

#### Case 1

A young boy presented with worsening headache and vomiting for 3 weeks and was admitted for further evaluation due to altered mental status. Imaging showed a 2.9 cm lesion in the right pons (Sample ID #1). Lumbar puncture and CSF studies were conducted, which revealed a hypercellular CSF with rare enlarged atypical cells worrisome for malignancy by cytomorphologic exam (**Fig. 2A**). Immunohistochemical (IHC) stains (GFAP, Neu-N, CD68) were ordered on the cell block; however, no atypical cells were identified. The cytology interpretation was limited due to the scant number of atypical cells seen in the cytospin preparations and a biopsy was recommended for a definitive diagnosis. Tumor tissue biopsy revealed a cellular specimen morphologically consistent with a high-grade glioma (**Fig. 2B**). Pertinent IHC stains suggested mutations in H3K27M (gain) and H3K27me3 (loss) (**Fig. 2C**), The final diagnosis was reported as diffuse midline glioma (DMG), H3 K27-altered, CNS WHO grade 4. MGMT gene promoter methylation was not detected. Due to the small size of the tumor tissue from a critical anatomical location, neither NGS could be performed on the tiny tissue sample nor surgical resection in this patient. The CSF cytology supernatant was collected, and LP-WGS was performed. In addition to other alterations, the results demonstrated a high copy number gain in chromosome 4, spanning positions 39,333,653 to 68,681,040, which includes the platelet-derived growth factor receptor alpha (*PDGFRA*) gene (**Fig. 2D-E**). *PDGFRA* is frequently amplified and mutated in pediatric high-grade gliomas, representing the most common target of focal amplification in these tumors, including DMG. Subsequently, a targeted brain tumor NGS panel was performed on ctDNA from the CSF supernatant. The results confirmed the *H3-3A* c.83A>T, p.K28M mutation with a variant allele frequency (VAF) of 12.5%. This finding aligns with the IHC results from tumor biopsy tissue. Additionally, the panel detected a TP53 gene mutation (c.722C>T, p.S241F) with 5% VAF.

**Figure 2:**
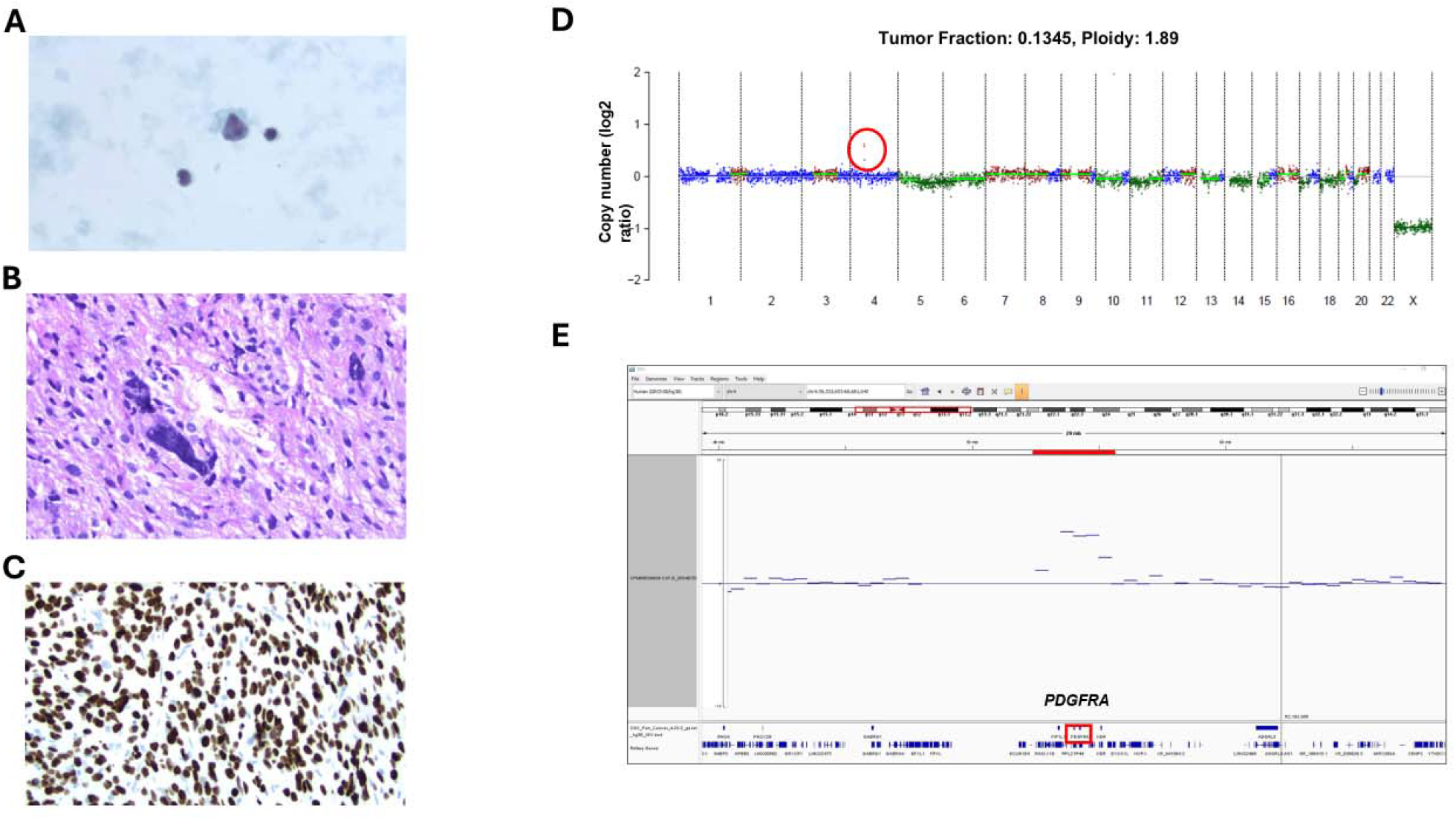
**(A)** CSF cytologic examination from Case 1 revealed rare, atypical cells. **(B)** Hematoxylin and Eosin stained (H&E) slides showed a cellular primitive tumor with marked cytologic anaplasia. **(C)** Immunohistochemistry (IHC) revealed H3K27M mutant protein with loss of expression of H3K27me3. **(D-E)** LP-WGS was performed on the CSF cytology supernatant from patient ID #1 (diffuse midline glioma). This revealed a copy number gain at chromosome 4 (red circle), spanning positions 39,333,653 to 68,681,040, which includes the *PDGFRA* gene.

#### Case 11

A previously healthy preschool-aged girl presented to an outside emergency department with a severe headache, left sided facial droop, left upper extremity weakness, tremors, and ataxia. CT head imaging demonstrated a 6.6 cm right fronto-parieto-temporal mass causing a 2 mm midline shift. One day after admission, the patient was noted to have unequal pupils, headache, emesis, and worsening left sided weakness. Magnetic resonance imaging (MRI) demonstrated a 6.3 cm large solid and cystic mass with a 15 mm leftward subfalcine herniation (**Fig. 3A**). The patient’s condition continued to decline, and she was subsequently taken to the operating room for mass removal. Post-surgical MRI showed residual tissue remaining in the tumor bed. Histologic examination of the tumor tissue demonstrated areas with primitive embryonal tumor, frequent multinucleated tumor cells with cytologic anaplasia, myxoid stroma with atypical mitotic figures, and fragments of tumor with low-grade appearance (**Fig. 3B**). Pertinent IHC stains demonstrated positivity for neurofilament protein (NFP) and oligodendrocyte transcription factor 2 (OLIG2) and high indexes for p53 (80%) and Ki-67 (80%). Molecular reports included the following genetic mutations: *H3-3A* mutant, K28M (K27M); *BRAF* V600E; *PDGFRA* R841-I843delinsKV; *TP53* R273C; *PD-L1* LDT. CNAs included chromosome 7 gain and equivocal MYCN amplification. No MGMT gene promoter methylation was detected. The final diagnosis was reported as DMG, H3 K27-altered, CNS WHO grade 4. Several weeks later, the patient presented for follow-up and MRI imaging which showed re-enhancement. Cytology of the CSF collected at this time revealed a cellular specimen with scattered atypical cells worrisome for malignancy (**Fig. 3C**). Reactive atypia could not be ruled out as the cells of interest were not identified on the cell block by IHC (glial fibrillary acidic protein [GFAP], synaptophysin). Supernatant from fresh CSF and cytology CSF samples were sent for analysis. The samples were negative for genomic variations by LP-WGS, but NGS demonstrated *BRAF* and *H3-3A* mutations. The results confirmed the *H3-3A* c.83A>T, p.K28M mutation, with a VAF of 24% in fresh CSF and 14% in the cytology supernatant. The *BRAF* V600E mutation was also detected, with a VAF of 27% in fresh CSF and 17% in the cytology supernatant. These findings, which were only reported by the NGS panel, aligned with the NGS results from the resected tumor tissue. Of note, NGS of the fresh supernatant reported an additional mutation, *CDKN2A* (VAF of 1%). Further CSF studies were not conducted as the tumor had progressed, and the patient was transitioned to hospice care.

**Figure 3:**
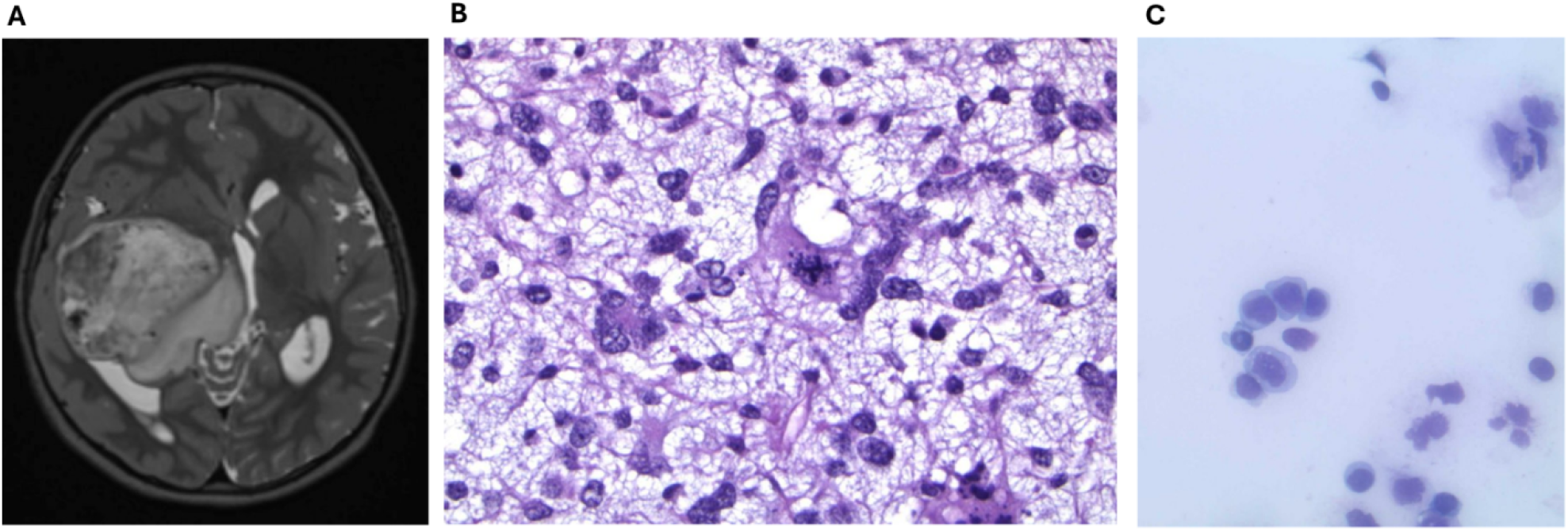
**(A)** MRI of the head from Case 11 demonstrated a 6.3 cm large solid and cystic mass with a 15 mm leftward subfalcine herniation. **(B)** H&E slide showing areas with primitive embryonal tumor with myxoid stroma and atypical mitotic figures. **(C)** CSF cytologic examination revealed a cellular specimen with scattered atypical cells.

### cfDNA sequencing from serial cytology supernatants can successfully monitor patient progression

#### Case 4

A previously healthy male toddler presented with congestion, coughing and emesis. He was diagnosed with croup and given supportive measures. However, despite initial improvement, the patient’s symptoms continued to worsen. CT imaging demonstrated hydrocephalus due to a 3.7 cm posterior fossa mass centered in the fourth ventricle, with extension through the right foramen of Luschka and into the cisterna magna and the patient was subsequently admitted to the pediatric intensive care unit (PICU). MRI demonstrated a large, heterogeneously enhancing lesion of the posterior fossa both dorsal to the cerebellar hemispheres and extending into and filling the fourth ventricle, and extending through the right foramen of Luschka towards the cerebellopontine angle and compressing the brainstem medially towards the left side (**Fig. 4F**). There was also evidence of diffuse leptomeningeal disease throughout the spine, consistent with drop metastasis. Cytology of the collected CSF revealed clusters of poorly differentiated tumor cells (**Fig. 4E**). The patient was started on induction chemotherapy, however end of induction scans demonstrated progression of disease. Repeat MRI scans continued to show slight increase in leptomeningeal disease. After continued treatment, the patient showed stable scans and was eligible for autologous transplant. Histologic examination demonstrated a high grade, poorly-differentiated tumor composed of small round monomorphic cells with scant cytoplasm and round nuclei that are hyperchromatic with delicate chromatin (**Fig. 4A**). Mitotic activity and apoptotic bodies were seen without frank necrosis. IHC stains were notable for synaptophysin with a dot-like perinuclear staining and high Ki-67 of 90% (**Fig. 4B-C**). All other markers were negative. FISH analysis for MYC and MYCN as well as C19MC did not detect amplification. The tumor was diagnosed as medulloblastoma, non-WNT/non-SHH, group 3/4, subgroup 4 (subgroup 4 of 8) by methylation profiling of tumor tissue DNA. NGS profiling of tumor tissue DNA revealed copy number losses of chromosome 4, 8, 9, 10, 11, and 21, with no chromosome gains. LP-WGS of fresh CSF was performed and revealed copy number losses of chromosomes 8, 9, 10, 11, 16, and 21, similar to the tumor tissue results. The LP-WGS also revealed copy number gains of chromosome 1, 2p, 3, 5, 7, and 14 which were not previously identified in the tumor tissue. NGS of fresh CSF revealed a *PDGFRA* c.2534A>G mutation with VAF of 69% (**Fig. 4D**).

**Figure 4.**
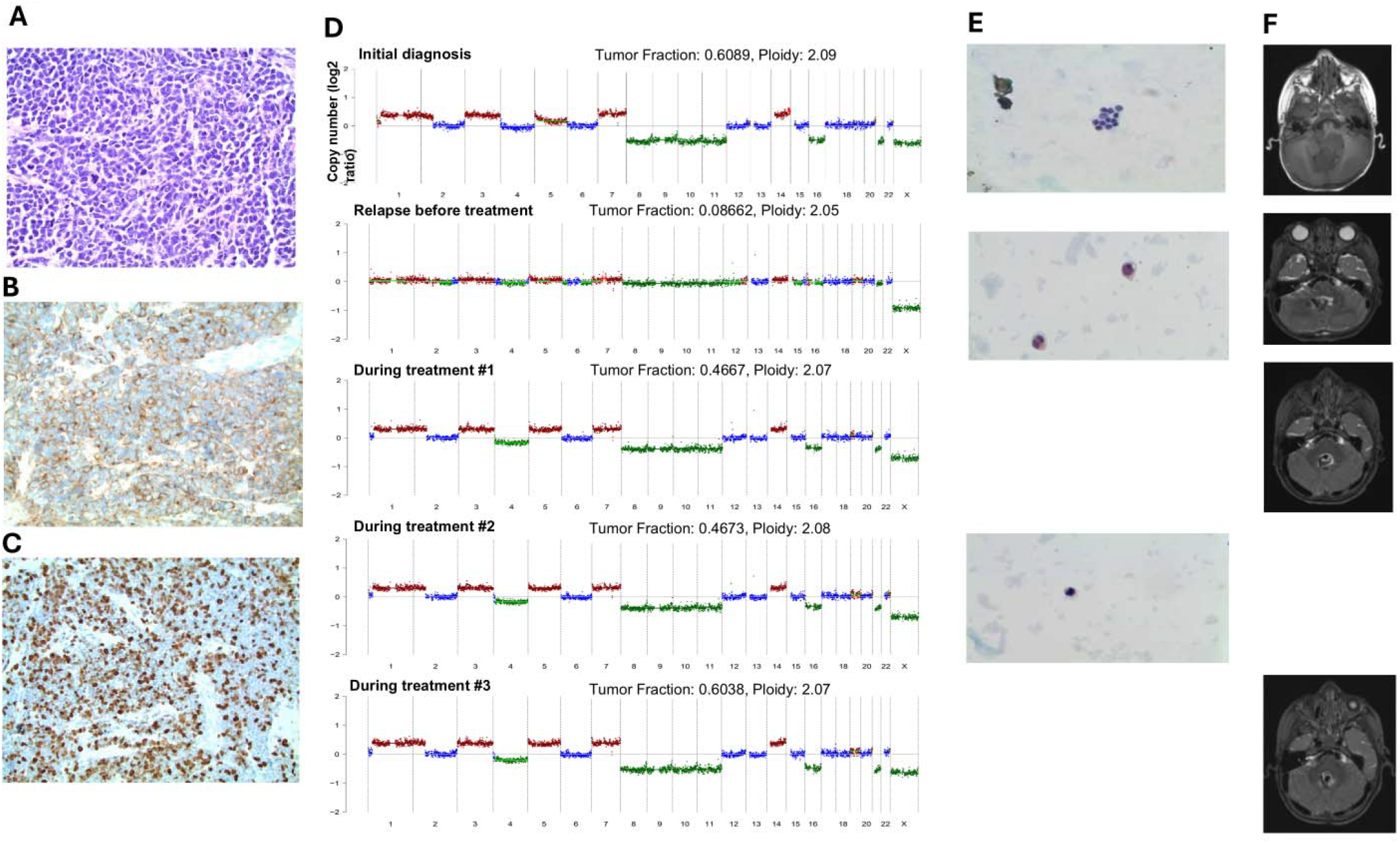
Tumor LP-WGS CNA, cytologic and radiographic analysis from Case 4, diagnosed with group 3/4 medulloblastoma. **(A)** H&E slides showed a high grade, poorly-differentiated tumor composed of small round monomorphic cells with scant cytoplasm and round nuclei that are hyperchromatic with delicate chromatin. **(B-C)** IHC stains were positive for synaptophysin with a dot-like perinuclear staining **(B)** and high Ki-67 of 90% **(C). (D)** LP-WGS was performed on the initial diagnosis and four follow up CSF cytology supernatants (relapse prior to MEMMAT protocol treatment, and during subsequent stages of treatment with treatment #1 occurring at day 17, treatment #2 occurring at day 40 and treatment #3 occurring at day 46 of MEMMAT protocol). **(E)** CSF cytologic examination results at the corresponding timepoints. **(F)** MRI results at the corresponding timepoints.

Four follow-up CSF cytology supernatants were collected through the shunt: at relapse before treatment and during three treatment phases (#1-3) but no malignant cells were identified (**Fig. 4E**). The samples demonstrated an additional chromosome 4 loss compared to the initial diagnosis **(Fig.4D** and **Table 1)**. The ctDNA fraction was low at the time of relapse but continued rising during treatment (**Supplementary Table 1**). Throughout treatment after relapse, MRI demonstrated stable tumor size (**Fig. 4F**). These findings were consistent with clinical treatment resistance. CSF cytologic examination was negative for malignant cells at relapse and throughout treatment (**Fig. 4E**).

#### Case 8

A teenaged boy with a history of achondroplasia, conductive hearing loss, and developmental delay presented with new onset seizures. A MRI of the head showed a 4.9 cm heterogeneously enhancing and diffusion restricting mass centered within the right cerebellum and extending to the right cerebellar vermis with associated regional vasogenic edema and mass effect (**Fig. 5A**). MRI of the L-spine demonstrated a 5.7 cm space-occupying intradural and extramedullary enhancing mass centered within the spinal canal and extending from approximately L4-S1 with resultant severe displacement of the thecal sac and mild bilateral neural foraminal narrowing (**Fig. 5B**). Histological findings demonstrated a small round blue cell malignancy expressing synaptophysin, beta catenin (cytoplasmic) and INI1 by IHC staining (**Fig. 5C**). NGS testing of the tumor tissue was positive for *FGFR3* G380R, *PTCH1* A451Gfs*6, *TERT* promoter c.-124C>T and *TP53* R175H. Tumor tissue CNAs included a gain of chromosome 7 gain and amplification of *EGFR. MYCN* amplification was noted but considered equivocal. No MGMT gene promotor methylation was identified by methylation profiling. The final diagnosis was reported as medulloblastoma, WHO grade 4 non-SHH, non-WNT. The patient underwent adjuvant therapy. Several months later, surveillance imaging demonstrated a small enhancing abnormality deep in the right frontal periventricular region. The CSF specimen examined prior to surgery showed no malignant cells. Immediate postoperative MRI demonstrated a significant interval growth in size of the tumor compared to the preoperative MRI with a new abnormal lesion in the genu of the corpus callosum, intraventricular and at the head of the caudate, concerning for rapid growth and suggestive of an incredibly aggressive tumor. A repeat craniotomy was done followed by the primary treatment. Unfortunately, as the disease progressed, the patient was transitioned to hospice care and passed away shortly thereafter.

**Figure 5:**
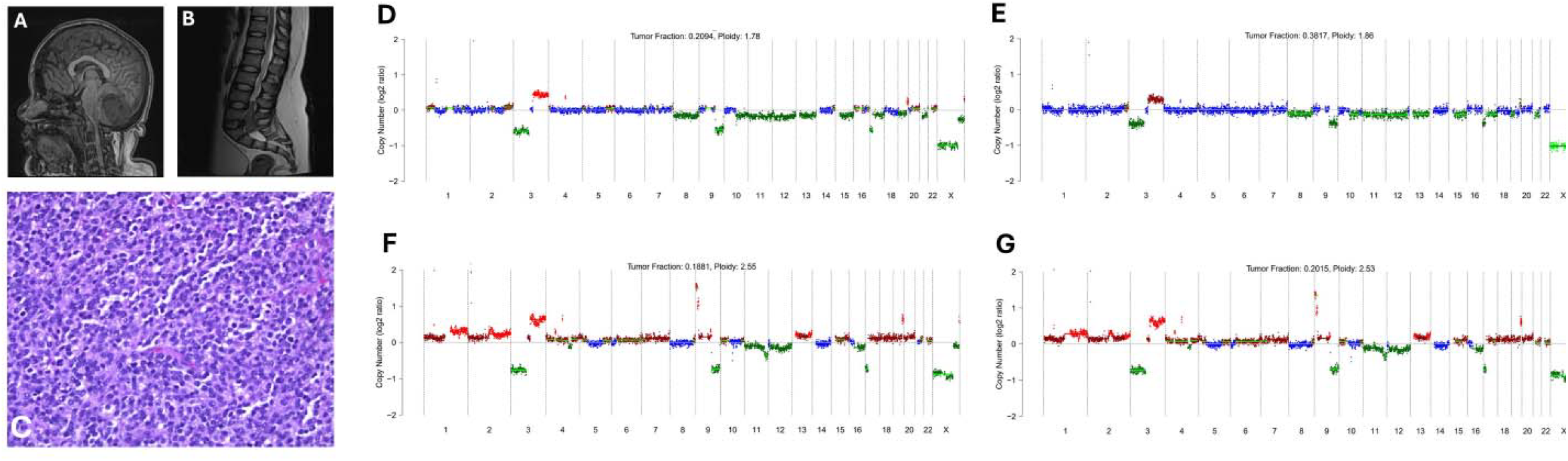
**(A)** MRI of the head demonstrated a 4.9 cm mass in the cerebellum, extending to the vermis. **(B)** MRI of the spine demonstrated a 5.7 cm intradural, extramedullary mass within the spinal canal from L4-S1. **(C)** H&E section of medulloblastoma, WHO grade 4. Baseline CSF of **(D)** fresh CSF and (E) cytology CSF LP-WGS showed a complex copy number profile, including *MYCN* amplification, 3p loss, 3q gain, loss of 8, 9q, 10q, 11, 12, 13, 15, 17p, loss of 19p and distal gain of 19, loss of 21 and partial gain of Xq. Follow up analysis of **(F)** fresh CSF and **(G)** cytology CSF LP-WGS showed a more complex copy number profile than the baseline CSF specimens, including new chromosome gains of different amplitudes in chromosomes 1 and 2, high level gains in 9p, deletion in 11q, gain of 13 and loss of 16.

The initial CSF specimen at relapse, which was benign on cytology, was sent for analysis. LP-WGS of the fresh CSF supernatant revealed a complex copy number profile, including *MYCN* amplification, 3p loss, 3q gain, loss of 8, 9q, 10q, 11,12, 13,15, 17p, 19q, gain of distal 19q, loss of 21 and gain of distal Xq **(Fig. 5D)**. The same results were reported for the LP-WGS of the cytology derived supernatant **(Fig. 5E)**. Targeted NGS of the fresh CSF supernatant revealed the following mutations: *TP53* c.524G>A, p.R175H (62%); FGFR3 c.1138G>A, p.G380R (46%); *PTCH1* c.1351_1352insGG, p.A451Gfs*6 (52%); *DDX3X* c.1615+1_1615+2dupGT (59%); and *TERT* c.-124C>T (40%). The NGS of the cytology supernatant demonstrated the same mutations but at lower VAF’s (**Table 1**). The mutations reported by NGS of the tumor tissue were reflected in that of the supernatant except for *DDX3X*. Approximately five months later the patient returned for follow up, and CSF was collected for cytology and molecular analysis. LP-WGS of the fresh CSF and cytology derived supernatants showed a more complex copy number profile than the baseline CSF specimens, including chromosome gains at different amplitudes in chromosomes 1 and 2, high level gains in 9p, deletion in 11q, gain of 13 and loss of 16 as well as the previously noted CNAs **(Fig. 5F)**. In addition to the mutations reported in the previous analysis, NGS of the fresh CSF supernatant identified two new mutations: *MYCN* c.134delC, p.P45Rfs*86 (0.2%) and *CREBBP* c.5214_5234del, p.H1738_W1745delinsQ (6%) **(Fig. 5G)**. The *MYCN* mutation, detected at a very low VAF, corroborated the findings from tissue-based NGS. This may reflect the emergence of a dominant tumor subpopulation potentially driving disease progression. Since the initial analysis, VAF’s of the established mutations in follow up specimens all increased.

## Discussion

In this study, we have shown that the CSF supernatant from cytology specimens can serve as valid liquid biopsy source for cfDNA sequencing. As shown in **Case 1**, the molecular characteristics of the tumor were identified in the CSF supernatant from the cytology specimen even when testing of subsequent tumor biopsies was not possible. From the supernatant, LP-WGS revealed *PDGFRA* gain, likely representative of the amplification seen in the primary tumor, and targeted NGS confirmed an *H3-3A* c.83A>T, p.K28M mutation. In addition, the NGS panel detected a *TP53* c.722C>T, p.S241F mutation (**Table 1**). In **Case 11**, the LP-WGS results for both the fresh CSF supernatant and cytology supernatant were negative. However, the targeted NGS panel revealed *BRAF* and *H3-3A* mutations (**Table 1**), highlighting the importance of a comprehensive detection platform.

In addition, we prospectively enrolled patients at initial diagnosis and to monitor disease progression. In **Case 4**, LP-WGS of the CSF cytology supernatant demonstrated copy number losses that were observed in the tumor tissue as well as copy number gains of chromosome 1, 2p, 3, 5, 7, and 14 that were not previously identified in tumor tissue, potentially representing a more accurate assessment of the subclonal composition of tumors (**Table 1**). Notably, gains of chromosome 14—particularly involving the *OTX2* gene—and chromosome 7 have been associated with medulloblastoma progression^13,14^. *OTX2* copy number gain is recognized as a molecular driver event in aggressive medulloblastoma subtypes^13^. NGS revealed a *PDGFRA* c.2534A>G (**Table 1**). In the subsequent follow-up evaluations, CSF samples had an additional chromosome 4 loss. Furthermore, the fraction of ctDNA in the CSF gradually increased during treatment which correlated with the clinically observed treatment resistance. In **Case 8**, NGS of the fresh CSF and cytology supernatant highlighted a *DDX3X* mutation which was not detected with NGS of the tumor tissue. In the follow-up liquid biopsy analysis, a *MYCN* mutation was detected in the fresh supernatant, which was not present in the initial supernatant analysis (**Table 1**). Identification of the *DDX3X* mutation may support the utility of comprehensive analysis of liquid biopsies as a better method for detecting tumor heterogeneity compared to standard molecular profiling assays. In addition, this demonstrates that many brain tumors are heterogenous in nature and additionally continue to acquire more mutations over time that may lead to tumor resistance, which has been noted in the literature in the past^15^. Although the *MYCN* mutation was not detected in the initial liquid biopsy specimen, detection in the follow up specimen during relapse presumably reflects the presence of the *MYCN* mutation in the residual tumor. Liquid biopsy analyses may be more sensitive in detection genomic evolution than tumor biopsies over time. In Cases 4 and 8, the increase in ctDNA fraction and the additional mutations detected by LP-WGS and the targeted NGS panel supports the findings from other studies that have demonstrated tumor heterogeneity and genomic evolution over time as revealed by liquid biopsy ^16,17^.

Comparison of the fresh CSF and cytology samples provided valuable insight when considering the use of cytology supernatants for liquid biopsy analysis. For example, the LP-WGS and targeted NGS panel of the fresh CSF and cytology supernatants detected the same gene mutations in Case 8. However, the VAF in the fresh supernatant was higher than that of the cytology supernatant (**Table 1**). In the follow-up liquid biopsy, there were several notable differences. First, the *MYCN* mutation was identified only in the fresh supernatant. Second, most of the genes in the fresh supernatant had higher VAFs except for *CREBBP* and *PTCH1*, which had the same VAFs (**Table 1**). Based on our limited series, there may be a quantitative difference between the ctDNA in fresh supernatant and cytology derived supernatant. The differences in VAFs between the two specimens may be attributed to cfDNA degradation occurring during the interval between collection and processing, likely due to the delay while awaiting completion of the CSF cytology examination. Immediately freezing the CSF cytology supernatant at −80°C may help to minimize degradation at room temperature.

Recent genomic and therapeutic advances have improved survival for pediatric patients with primary CNS tumors. However, the outcomes for high grade CNS tumors are still poor, especially after tumor recurrence. Early detection of markers such as MYC upon recurrence provides information about treatment resistance and highlights the need for better treatment approaches. Current standard practice for monitoring disease recurrence and risk stratification relies on imperfect tools. These include MRI, which is insufficient for detecting microscopic tumor cells, and CSF cytologic examination, which is often non-diagnostic because of paucicellular CSF. In addition, the surgical biopsies are invasive, making serial tissue biopsies extremely difficult to perform. There is a pressing need for a reliable and minimally invasive tool to monitor disease status, improve risk stratification, and guide treatment regimens. Recent studies have shown that LP-WGS can detect measurable residual disease (MRD) through identification CNAs before radiographic evidence in medulloblastoma^18^. O’Halloran et al. performed LP-WGS on serial CSF liquid biopsies from patients with recurrent medulloblastoma, revealing a correlation between the presence of CNAs with tumor evolution and treatment response^17^. These findings highlight the prognostic significance of CSF liquid biopsy in monitoring disease. However, multi-omic biomarkers in serial CSF cytology supernatant-derived ctDNA for primary pediatric CNS tumors have not been investigated. In our study, Case 3 and 5 yielded negative results by both LP-WGS and the targeted NGS panel. Case 3 was a child with a diagnosis of H3 K27-altered diffuse midline glioma. Case 5 was a teenager who initially presented with pilocytic astrocytoma with “oligodendroglioma-like features” with presumed progression to diffuse pediatric-type high grade glioma. In both cases, neither the LP-WGS nor the targeted NGS panel demonstrated alterations suggested by the clinical presentation or prior tumor tissue testing. Whether this was due to limited sampling or other biologic factors is not known.

In summary, we observed that combined LP-WGS and targeted NGS can detect ctDNA in the CSF cytology supernatant samples, monitor ctDNA fraction, and demonstrate tumor heterogeneity and genomic evolution over time. By utilizing routinely collected cytology specimens, our approach transforms clinically discarded specimens into a reliable source for liquid biopsy. CNAs and genetic mutations detected in CSF-derived cfDNA from patients with high grade glioma and embryonal tumors can augment tissue-derived molecular analyses. CSF liquid biopsy has the potential to transform clinical practice for improved diagnosis, risk stratification, and disease surveillance. Future research will be necessary to optimize methodology and enable clinical implementation.

## Acknowledgments

We appreciate the Cytology Laboratory, Department of Pathology and Human Anatomy at Loma Linda University, Loma Linda, California, for their assistance with CSF cytology processing. We appreciate the support provided by Dr. Paul Herrmann, Chairman of the Department of Pathology and Human Anatomy.

## Ethics

Loma Linda University institutional review board approved the project.

## Authorship statement

Y.C.W conceived the research and supervised all aspects of the work. J.B oversaw all aspects of liquid biopsy platform and laboratory activities. N.C, E.C and B.E drafted the manuscript. M.B, R.R, T.M, D.K.T, M.S and B.O enrolled patients and collected samples. N.C, E.C, K.L, B.F and Y.C.W processed samples. J.B, L.K, Y.C.W, R.N, I.K, G.Y, G.O and C.F contributed to data analysis. R.R, U.O, P.S, and A.A served as consultant. All the authors read, edited, and approved the manuscript.

## Data Availability

De-identified data will be made available upon reasonable request.

---

**Supplementary Table 1.** Fresh versus cytology supernatant CSF yields.

| Case | Source | CSF collect methods | Total cfDNA concentration (ng/ul) | ctDNA fraction (%) | Days between collection and freeze |
| --- | --- | --- | --- | --- | --- |
| 1 | Cytology | LP | 0.259 | 0.1345 | 2 |
| 3 | Fresh | EVD | 1.63 | 0.1365 | 0 |
|  | Cytology | EVD | 0.471 | 0.0632 | 5 |
| 4 | Fresh | EVD | 8.1 | 0.6089 | 0 |
| 4 1<sup>st</sup> f/u | Cytology | LP | 0.345 | 0.0866 | 10 |
| 4 2<sup>nd</sup> f/u | Fresh simulate cytology | Intrathecal chemotherapy VP shunt | 0.129 | 0.4667 | 0 |
| 4 3<sup>rd</sup> f/u | Cytology | LP | 0.0914 | 0.4673 | 6 |
| 4 4<sup>th</sup> f/u | Fresh simulate cytology | Intrathecal chemotherapy VP shunt | 0.129 | 0.6038 | 0 |
| 5 | Fresh | EVD | 2.92 | 0.0842 | 0 |
|  | Cytology | EVD | 1.42 | 0.0663 | 6 |
| 6 | Cytology | LP | 0.0853 | 0.0346 | 5 |
| 8 | Fresh | LP | 0.001 | 0.2094 | 0 |
|  | Cytology | LP | 0.0385 | 0.3817 | 3 |
| 8 f/u | Fresh | VP shunt tap | 0.116 | 0.1881 | 0 |
|  | Cytology | VP shunt tap | 0.0504 | 0.2015 | 7 |
| 11 | Fresh | LP | 2.04 | 0.0851 | 0 |
|  | Cytology | LP | 2 | 0.0787 | 26 |

EVD: external ventricular drain; LP: lumbar puncture; VP shunt: ventriculoperitoneal shunt.

